# Using simulated infectious disease outbreaks to guide the design of individually randomized vaccine trials

**DOI:** 10.1101/2021.01.29.21250703

**Authors:** Zachary J. Madewell, Ana Pastore Y Piontti, Qian Zhang, Nathan Burton, Yang Yang, Ira M. Longini, M. Elizabeth Halloran, Alessandro Vespignani, Natalie E. Dean

## Abstract

**Background/Aims:** Novel strategies are needed to make vaccine efficacy trials more robust given the uncertain epidemiology of outbreaks. Spatially resolved mathematical and statistical models can help investigators identify sites at highest risk of future transmission and prioritize these for inclusion in trials. Models can also characterize the uncertainty in whether transmission will occur at a site, and how nearby or connected sites may have correlated outcomes. A structure is needed for how trials can use models to address key design questions, including how to prioritize sites, the optimal number of sites, and how to allocate participants across sites.

**Methods:** We illustrate the added value of models using the motivating example of Zika vaccine trial planning during the 2015-2017 Zika epidemic. We used a stochastic, spatially resolved, agent-based transmission model (GLEAM) to generate 1,142 epidemics and site-level incidence at 100 high-risk sites in the Americas. We considered several strategies for prioritizing sites (average site-level incidence of infection across epidemics, median incidence, probability of exceeding 1% incidence), selecting the number of sites, and allocating sample size across sites (equal enrollment, proportional to average incidence, proportional to rank). To evaluate each design, we stochastically simulated trials in each hypothetical epidemic by drawing observed cases from site-level incidence data.

**Results:** When constraining the overall trial sample size, the optimal number of sites represents a balance between prioritizing highest-risk sites and having enough sites to reduce the chance of observing too few endpoints. The optimal number of sites remained roughly constant despite varying the targeted number of events, although it is necessary to increase the total sample size to achieve the desired power. Though different ranking strategies returned different site orders, they performed similarly with respect to trial power. Instead of enrolling participants equally from each site, investigators can allocate participants proportional to projected incidence, though this did not provide an advantage in our example because the top sites had a roughly similar risk profile. Sites from the same geographic region may have similar outcomes, so optimal combinations of sites may be those that are more geographically dispersed, even when these are not the highest ranked sites.

**Conclusions:** Mathematical and statistical models may assist in the design of successful vaccination trials by capturing uncertainty and correlation in future transmission. Although many factors affect site selection, such as logistical feasibility, models can help investigators optimize site selection and the number and size of participating sites.

## Background

To observe enough events to reliably measure the efficacy of a vaccine, phase III trials often enroll thousands or tens of thousands of participants across multiple sites. For endemic diseases like rotavirus or malaria, incidence may be low but is relatively predictable. Investigators can use historical data to guide the selection of trial populations assuming that future trends will be similar. Where incidence is lower than expected during the trial, investigators can expand the sample size at existing sites or increase participant follow-up to compensate. This strategy is unlikely to work for outbreak pathogens. Historical data may be only weakly predictive of future incidence at a location. In fact, for pathogens with high attack rates, an area with a large prior outbreak may be less susceptible to a subsequent outbreak if there is a build-up of population immunity. Alternatively, that area may be more prone to another outbreak if immunity wanes or the number of susceptible individuals is replenished. The outbreaks themselves are highly unpredictable – when and where they will occur, how many will become infected, and how long they will last. The 2014-2016 West African Ebola epidemic was emblematic of this challenge, with a Phase III trial in Liberia enrolling over 8,000 individuals but observing no events because the local outbreak subsided.^1^ In this situation, expanding enrollment at existing sites or extending follow-up of participants would not be able to compensate.

Novel strategies are needed to make vaccine trials more robust to the uncertain epidemiology of outbreaks.^2^ One recommended approach is to enable the addition of new sites over time using a master or core protocol framework.^3^ If transmission in early hotspots is brought under control before the study has reached a conclusion, the trial can continue at new hotspots. If the outbreak is declared over, the trial can be paused until a subsequent outbreak. Spatially resolved mathematical and statistical forecast models can assist investigators in selecting participating sites. Models can incorporate site-specific features such as population size and density, socioeconomic vulnerability, sociocultural acceptance, logistic feasibility, prior immunity estimated from traditional surveillance or serosurveys, ongoing local transmission, or risk of importation. For vector-borne diseases, models can capture vector presence or abundance, sensitivity to temperature and humidity, the spread of other diseases by the same vector, and whether other diseases interfere with the disease of interest. By integrating diverse data sources, models can help investigators identify sites at highest risk of future transmission and prioritize these for inclusion in the trial.^4^

Another advantage of simulation models for infectious disease trials is that they enable investigators to explore a range of trial design features.^5^ Projected incidence is important, but so is the uncertainty around that projection, including the probability of no or little future spread. When there is a chance that sites will have little or no transmission, it becomes more important to include multiple, geographically dispersed sites, to distribute this risk.

We illustrate the potential role of forecast modeling by using simulation data from a stochastic, highly spatially resolved, agent-based Zika virus (ZIKV) model^6^ that was used to inform Zika vaccine trial planning in 2016.^7^ Although the Zika epidemic subsided so that vaccine efficacy trials were not possible,^8^ these are the type of data investigators would have at their disposal when designing future efficacy trials for other infectious diseases. In addition to generalizable findings, we provide a plan for how future trials may analyze their modeling results to prioritize test sites, site size, and the total number of sites. We explore how disease models can be used to address key trial design questions, including how to rank sites, the optimal number of sites to include, and how to allocate participants across sites. Simulations can also be used to explore trial feasibility given financial, logistical, or time constraints. We further consider how correlation between sites due to geographic proximity or human movement impacts trial power.

### Simulation structure

#### Model

We used the Global Epidemic and Mobility model (GLEAM) to identify the top 100 sites in the Americas with the highest projected ZIKV probability of transmission and infection rates in 2017. These projections were prepared in 2016, reflecting the type of data available to investigators planning trials.

GLEAM, which has been described elsewhere,^6, 9^ is a discrete stochastic epidemic computational model incorporating high-resolution demographic, socioeconomic, temperature, vector occurrence probability, and human mobility data. The projections were calculated using discrete time steps of one day to simulate transmission dynamics, but the results are summarized as number of infections per month. The resulting dataset included the site name, population size, and number of simulated infections (both symptomatic and asymptomatic) by month for 1,142 simulated epidemics from January through December 2017 (Table S1). Population sizes for sites included all ages. Thus, we can examine both the range of projections for each site, as well as look across sites within an epidemic.

#### Trial design

We describe the design of a hypothetical individually randomized Zika vaccine efficacy trial. The primary outcome is total number of confirmed symptomatic ZIKV cases. Given a set of selected sites and a fixed enrolled population for each site, for which we consider various different combinations, we simulate a trial as follows. First, we select one of the 1,142 simulated epidemics, which has an associated annual infection attack rate for each site. We simulate the number of infected trial participants at each site as a binomial draw with the probability of infection set at the site-level attack rate, and then we draw the number of these with symptomatic disease assuming 20% symptomatic proportion.^10^ This yields the total number of cases at each site, which is then added across sites. We repeat the binomial draws 50 times at each site, and then across all 1,142 epidemics to generate 57,100 simulated trials.

Approximately 60 symptomatic infection events are needed to have 90% power to reject the null hypothesis that the vaccine efficacy ≤ 30% when it is actually 70% using a 1:1 allocation to vaccine or placebo. We therefore defined a successful (i.e., adequately powered) trial as finding ≥60 cases across all sites in one year; we also explored trial designs targeting 50 to 150 events.

In a sensitivity analysis, we consider the feasibility of trials when attack rates are uniformly lower than projected by the model. To explore this scenario, we restrict analyses to the 25% of simulated epidemics with lowest overall infection attack rates across all sites.

## Findings

### Number of sites

The first key design choice is the number of participating sites. Sites are ranked by mean incidence of infection across all simulated epidemics (Figure S1), and we consider designs including the top site, the top two sites, and so on. For this example, we constrain the overall sample size at 15,000 participants and allocate these participants equally across selected sites. We plot the distribution of the simulated number of cases for each design in Figure 1. Starting on the left side of the figure, the bimodal nature of outbreaks is apparent when five or less sites are included. While the median number of cases of the one-site design is highest relative to other designs, with a high upper tail observed for large outbreaks, there is notable mass near zero cases, when little transmission occurs at the site. As the number of sites increases, this bimodal phenomenon disappears; the probability of having zero cases decreases, but the median expected number of cases also decreases because lower incidence sites are included.

**Figure 1.**
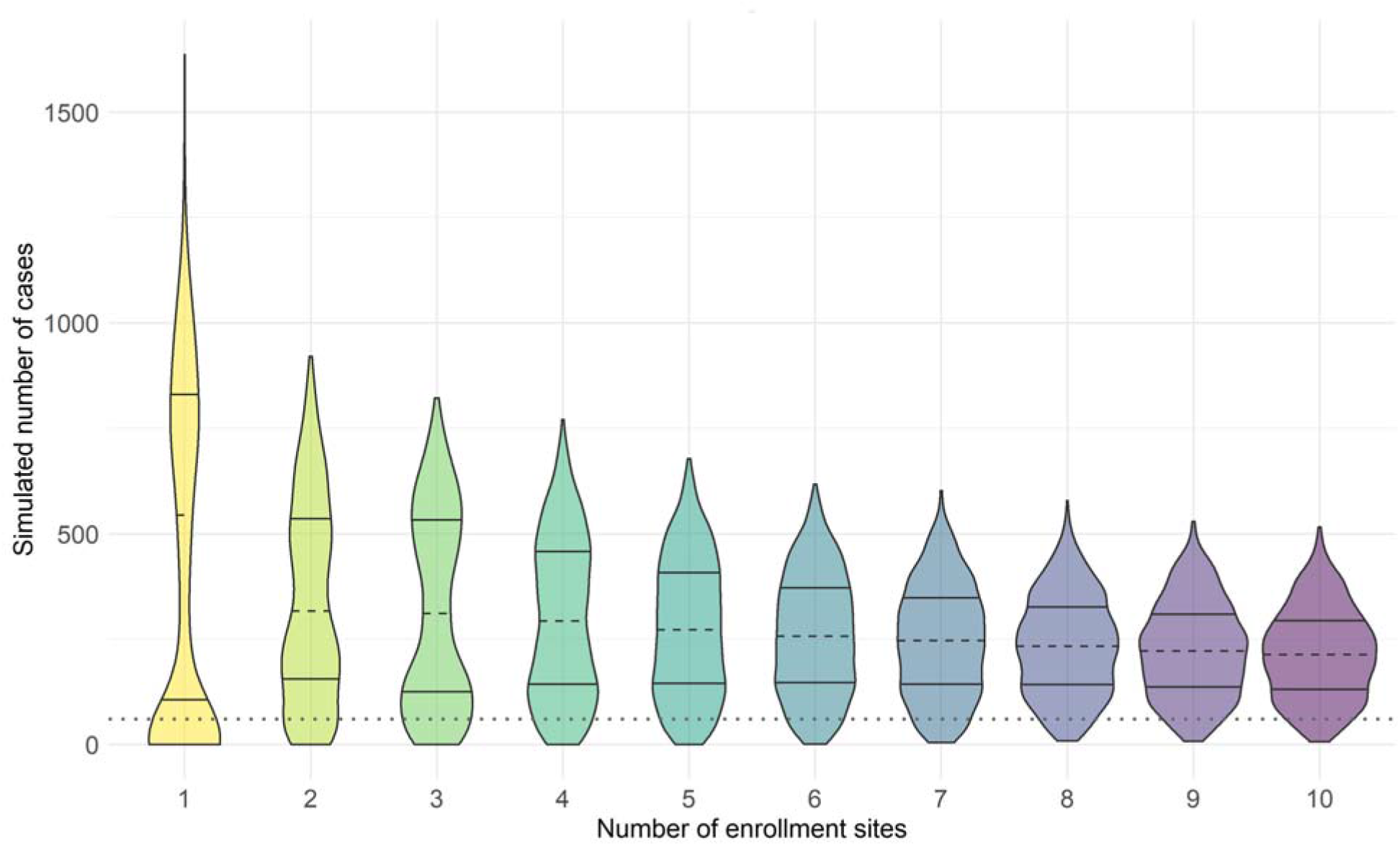
Violin plot of the simulated number of Zika virus cases for the top 1-10 sites with the highest average site-level incidence of infection across all simulated outbreaks in one year (2017). We assume an enrolled population of 15,000 across all enrollment sites with enrollment size spread evenly across all sites. Median number of cases (dashed line), 25^th^ and 75^th^ percentiles (solid lines) are shown. The threshold for a successful trial, defined as ≥60 cases across all sites in one year, is indicated by the dotted line.

While it is theoretically possible to enroll from only a single site, this presents an unacceptable risk of failing to accrue the needed endpoints. It may also not be practically feasible if the site has a small population. Furthermore, while a very high attack rate in a trial could shorten the trial duration or increase study precision, our primary goal is to meet our target number of events, not dramatically exceed it. Thus, rather than median expected number of cases, it is preferable to examine the probability that the design is adequately powered. The curves in Figure 2 A plot the probability of success (here defined as exceeding the target of 60 cases) as a function of the number of sites. We observe a local maximum around 8-11 sites, such that too few or too many sites are suboptimal with respect to trial success. In practice, the exact location of this maximum will depend on the specific epidemiological setting.

**Figure 2.**
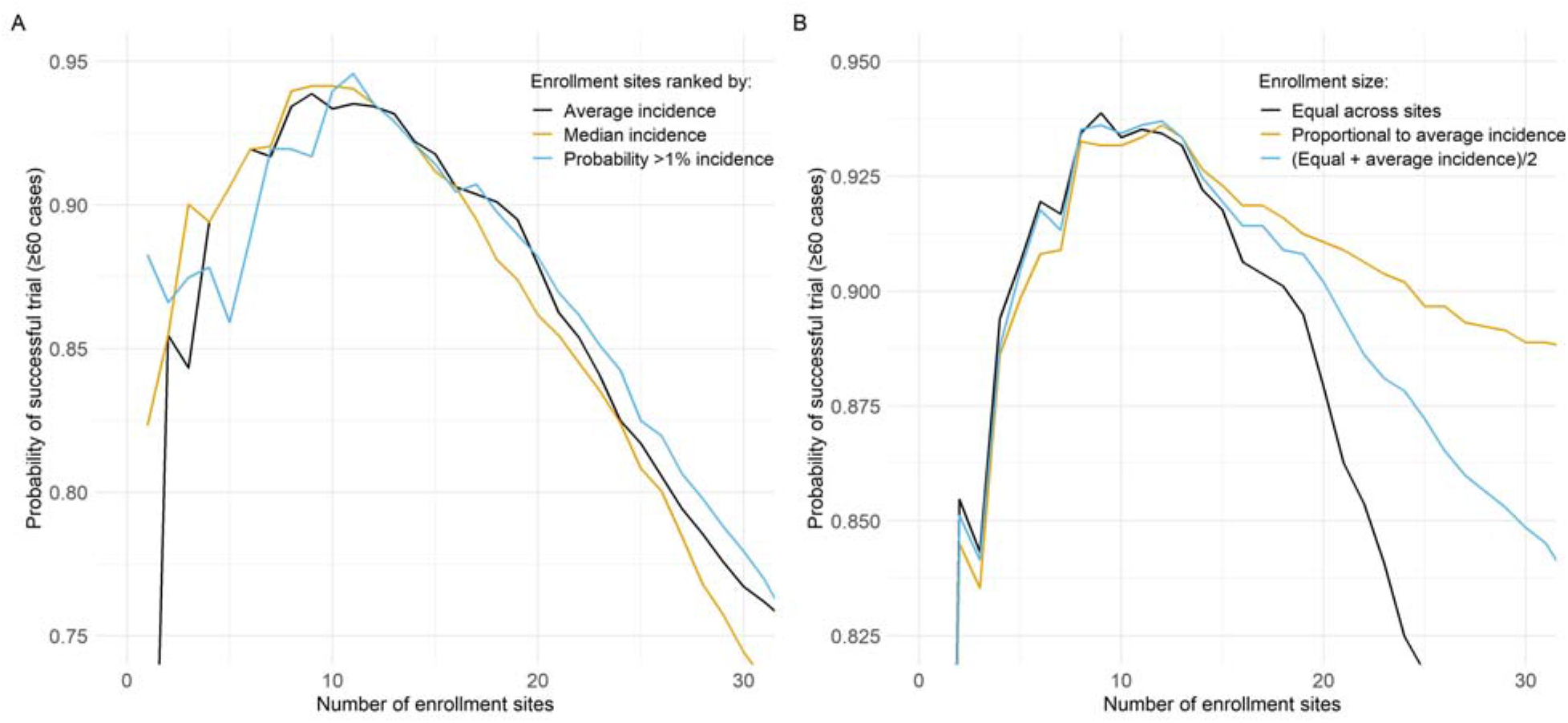
Probability of a successful trial (defined as ≥60 cases for an enrolled population of 15,000 across all enrollment sites in one year [2017]) as function of the cumulative number of enrollment sites. In Panel A, enrollment size was spread evenly across all sites and sites were added sequentially based on their ranking by the 1) average site-level incidence, 2) median incidence, and 3) probability of exceeding 1% site-level incidence of infection across all simulated outbreaks. In Panel B, sites were added sequentially based on their ranking of average site-level incidence of infection across all simulated outbreaks and enrollment size was 1) spread evenly across all sites, 2) proportional to the average site-level incidence of each site, and 3) average of equal enrollment and proportional to mean incidence.

The location of this local maximum for number of sites is reasonably stable to the ranking criterion, even if we change the target number of events or total sample size. Figure 3 shows the minimum trial size required to achieve at least 90% probability of success for different target numbers of events and sites. As the target number of events increases, there is an expected increase in the total number of participants needed. Yet for any specified target number of events, the desired 90% probability of success is achieved with the smallest overall sample size when around eight sites are included.

**Figure 3.**
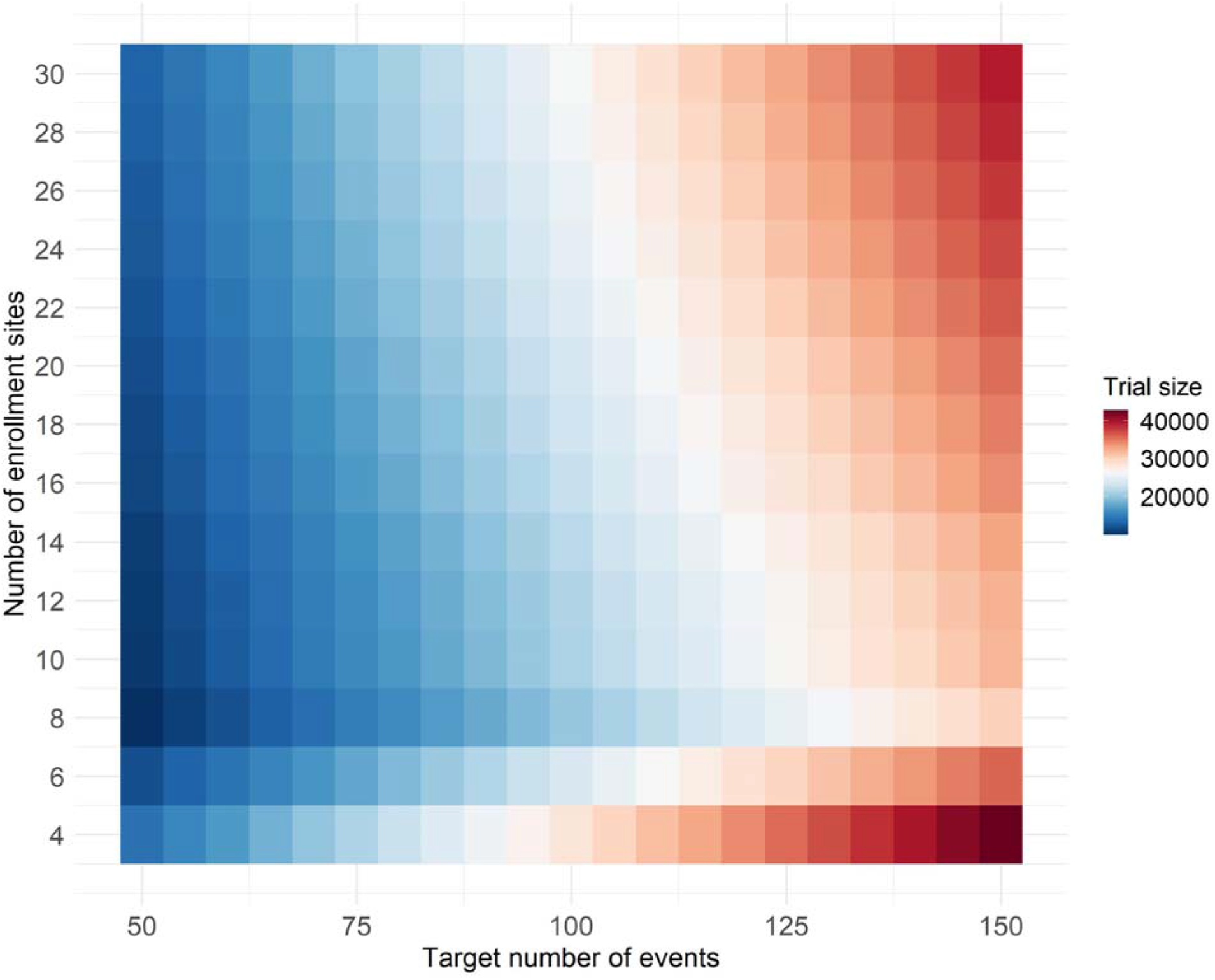
The minimum enrollment size required for an individually randomized vaccine efficacy trial to achieve at least 90% probability of success for different target numbers of events and numbers of sites. Sites were ranked by average site-level incidence of infection across all simulated outbreaks in one year (2017) with enrollment proportional to average site-level incidence of infection.

The optimal design depends on the underlying simulation data through the site-level attack rates. We consider a sensitivity analysis where the overall epidemic is smaller than projected by restricting to the 25% of simulations with lowest overall infection attack rates across all sites. Figure 4 compares the probability of success of different designs for all simulations versus the low incidence subset. While many of the same relationships persists, the probability of success drops dramatically. Even increasing the total number of sites and overall sample size, the probability of success does not exceed 25% in the designs explored. Thus, this approach is also useful for exploring the feasibility of trials.

**Figure 4.**
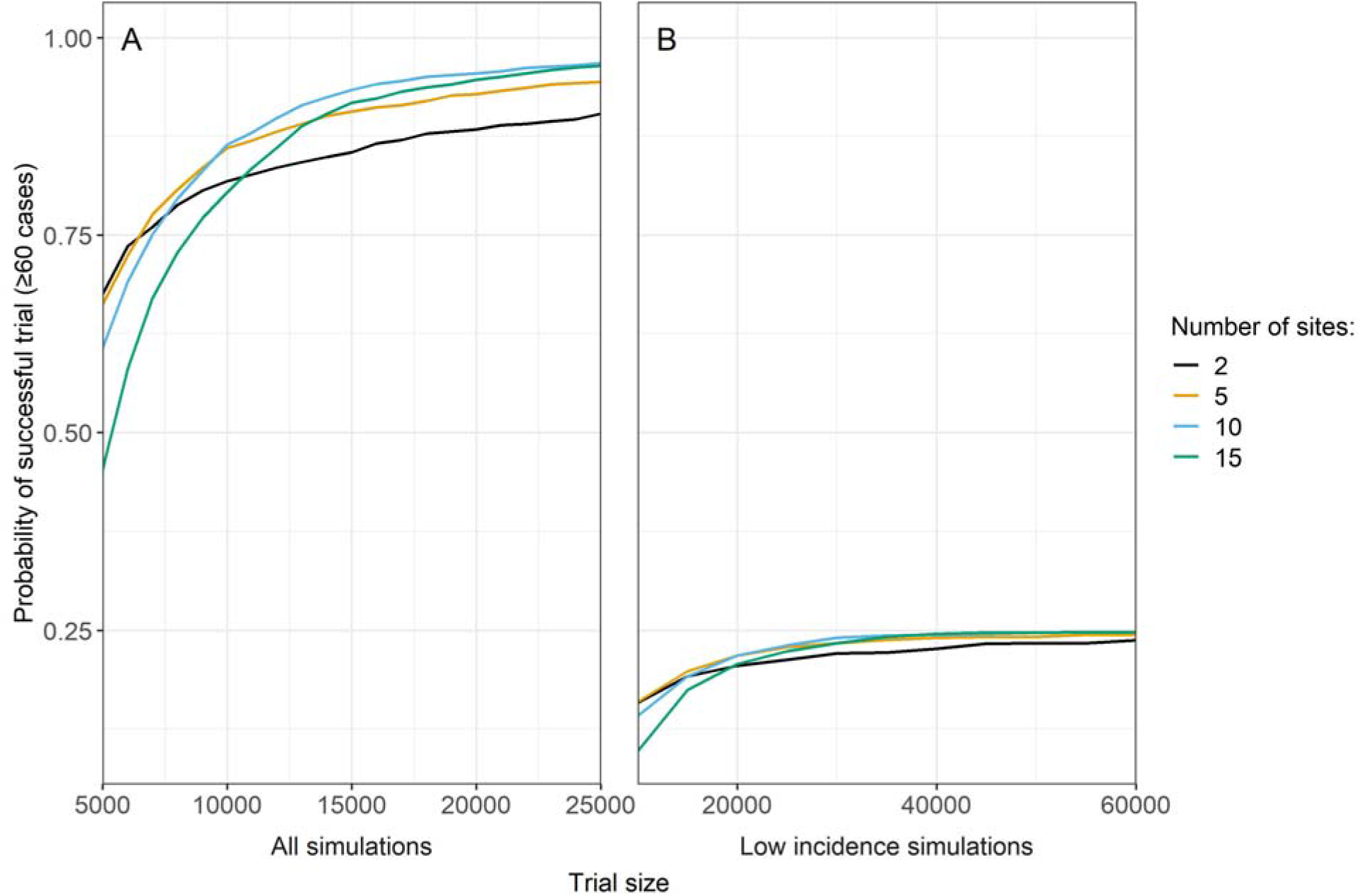
Probability of a successful trial (defined as ≥60 cases) as function of trial size, with enrollment size spread evenly across all sites. Sites were added sequentially based on their ranking by average site-level incidence of infection across all simulated outbreaks in one year (2017). Panel A includes all simulations, whereas Panel B is restricted to the 25% of simulated epidemics with lowest overall incidence across all sites.

Figure 4 also demonstrates that the two-site design is best when the targeted trial size is relatively small. Even for large targeted trial sizes, the difference in success probability between the two-site design and more sites designs is no more than 10%. Logistically, the two-site design is an appealing option.

### Site prioritization

In the previous section, sites are ranked by average model-projected site-level incidence of infection. We examined other ranking strategies, including median model projected site-level incidence, and the proportion of simulated outbreaks where site-level incidence exceeds a threshold, such as 1%. The latter strategy is intended to capture the bimodal nature of outbreaks, and that a few very large outbreaks could drive a high average incidence. In general, these measures are well-correlated, but they can yield different rankings (Figure S2, Table S2). Small sites may have higher attack rates, but may also have a higher probability of having zero infections across all participants. Nonetheless, we found similar performance across the different ranking strategies (Figure 2).

### Allocation strategies

Next, we considered different strategies for allocating the total sample size across multiple sites. The strategies are to distribute enrollment size 1) evenly across all sites, 2) proportional to mean incidence of infection, and 3) a middle-ground strategy using the average of sample sizes obtained from the previous two strategies.

1. Equal enrollment: Let *N* be the total sample size, let *m* be the number of sites, let *n*_*i*_ be the sample size at site *i*, then: 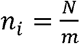
2. Proportional to mean incidence: Let *r*_*i*_ be the rank at site *i* and let *y*_*i*_ = mean incidence. Then site *i* has size: 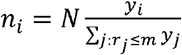
3. Average of equal enrollment and proportional to mean incidence: 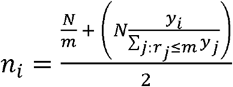

An example with five sites is shown in Table S3. The difference between the strategies will depend upon how similar projected incidence is across the top-ranked sites.

In this example, enrolling participants proportional to average incidence did not outperform the other strategies when fewer than 15 sites were included (Figure 2 B). This covers the range where the probability of success maximizes, around 8-11 sites. With larger numbers of enrollment sites, enrolling participants proportional to average incidence outperformed the other strategies as fewer individuals are enrolled from sites that are expected to have lower attack rates. However, designs with more sites are sub-optimal based on their reduced probability of success. As expected, the middle-ground strategy performs in between the others, but it may be desirable for logistical reasons by balancing enrollment across sites. The similar performance across the strategies for fewer than 15 sites may reflect that sites had similar enough risk and large enough uncertainty that proportional allocation provided no worthwhile advantage.

### Correlation between sites

It is conceivable that incidence rates are similar among sites in the same geographic region. Aside from Brazil, Figure 5 shows that correlation in incidence is highest among sites from the same country. It may therefore be that the sites with the highest simulated incidence are all from the same geographic region. To reduce the chance of enrolling sites from only one geographic area that may, by chance, have a smaller than expected outbreak, it may be prudent to simultaneously enroll participants from other geographically dispersed sites.

**Figure 5.**
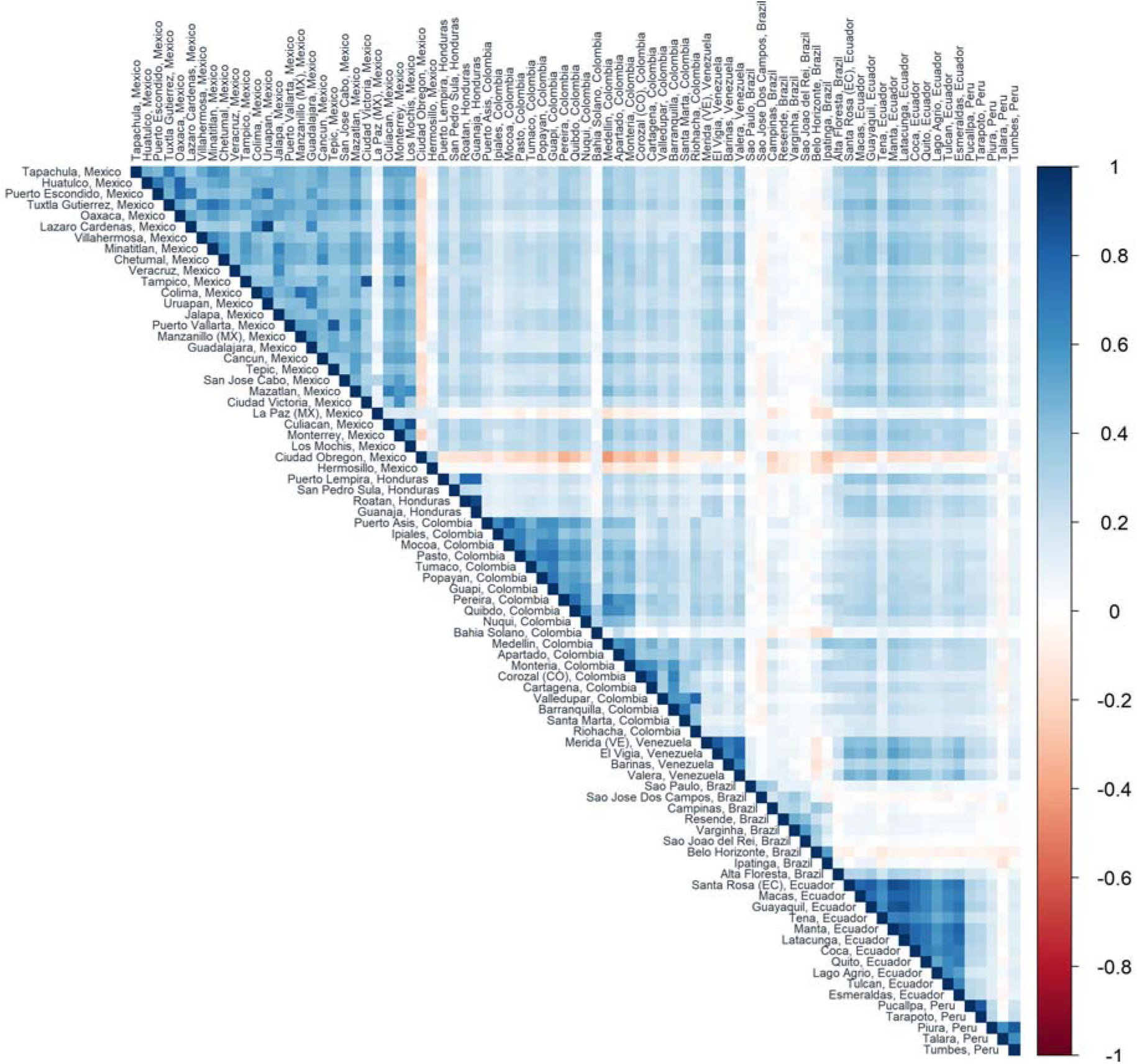
Spearman correlation of average site-level incidence of infection across all simulated outbreaks in one year (2017) between sites identified by the global epidemic and mobility model (GLEAM) (countries with at least four sites included). Sites are sorted by country and then by latitude.

We explored whether alternative combinations of sites (that may be more geographically dispersed) could have a higher probability of success than those based only on rankings. Figure 6 displays combinations of three, four, five, six, and seven site trials that achieved higher probability of success than a design that selects sites based solely on average site-level incidence. For example, the top three sites based on incidence are relatively close together, as seen by the low mean pairwise distance between sites plotted on the X-axis. Many other combinations of three sites return higher probability of success, and these tend to be more geographically dispersed (higher mean pairwise distance). A similar pattern is observed for higher numbers of sites, although the gains become more modest.

**Figure 6.**
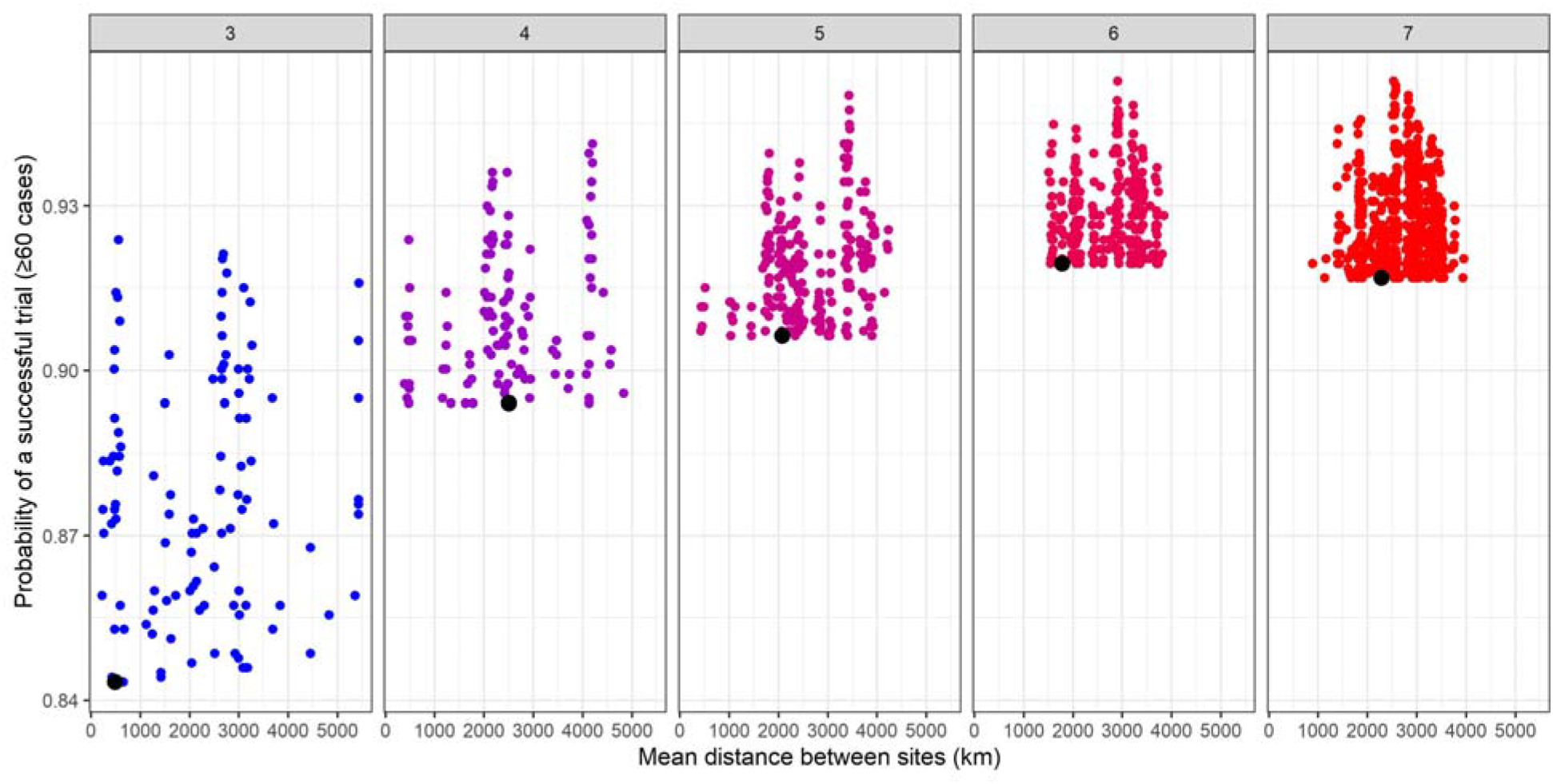
Probability of a successful trial (defined as ≥60 cases for an enrolled population of 15,000 across all enrollment sites in one year) by the mean distance between sites (in kilometers). The panels represent numbers of sites and points represent combinations of sites from the top 15 sites with the highest average site-level incidence of infection across all simulated outbreaks in one year (2017) that had a higher probability of success than the combination of sites with the highest projected incidence (represented by the black dot). Enrollment is assumed to be spread evenly across sites. We have included combinations of 3-7 sites of the top 15 sites for illustration, but this process could be extended to any number of sites.

### Number of countries

While it may be prudent to recruit sites from at least a few different countries (under the intuition that sites within a country are correlated), an important operational consideration is the number of countries enrolled. For each country included, the logistical burden for the trial increases substantially because it involves engaging with multiple ministries of health and the protocol needs to be approved by country-level institutional review boards. Thus, investigators may prefer to pursue trials in countries that have many high-risk sites. Figure S3 visualizes the number and incidence of sites by country. For example, in this scenario, it may be practical to select several sites from Peru, Mexico, Ecuador, and Colombia since many candidate sites are high risk.

## Conclusions

We describe the use of simulation data for design elements for individually randomized vaccine trials during an ongoing epidemic. Mathematical models allow us to capture a range of possible outcomes, from small to large outbreaks, and incorporate correlation between sites connected by human movement. Models therefore may capture the stochasticity of future transmission, reflecting a distribution rather than only the mean projection, which may be used to guide trial planning.

We used a single model to identify the top 100 sites in the Americas with the highest projected Zika virus transmission probability and infection rates in 2017 and leveraged those data to analyze vaccine efficacy trial design strategies. However, in a real-world infectious disease outbreak, ensemble forecast modeling (combining projections from independent modeling groups) may be used to expeditiously guide identification of appropriate sites for vaccine efficacy trials.^4^ In practice, implementation will depend upon practical considerations with regard to site selection, speed of rollout, number of participants across sites, including a site’s capacity to support a vaccine trial. There may also be political considerations.

The optimal number of sites to enroll represents a balance between having enough sites to distribute the risk of having a smaller than expected outbreak at any one site, versus enrolling participants from lower risk sites. This optimal number of sites stayed relatively constant even when increasing the targeted number of events, although the sample size must increase to achieve the desired probability of success. Though different methods of prioritizing sites (average incidence, median incidence, and probability of exceeding a threshold) returned different rankings, overall the approaches performed similarly in this example. In general, they return the same set of prioritized sites. Investigators can use simulations to guide allocation of participants across sites, prioritizing high-risk sites, though this appears to provide an advantage over a simpler equal allocation approach only when there are large differences in risk across sites.

We demonstrated that sites from a geographic region may have similar outcomes and that optimal combinations of sites may be those that are geographically dispersed. Perhaps it is feasible to develop novel search algorithms in the enormous space of site combinations without simulating trials for all possible combinations. However, including multiple countries increases trial costs and logistical complexities. Cost-effectiveness analyses may be used to explore the potential benefits and feasibility of specific trial design features by assessing the net costs or savings of vaccinating specific populations.^11^ Although many factors affect site selection, models can help investigators optimize site selection and the number and size of participating sites.

## Data Availability

The datasets analyzed during the current study are available from the corresponding author on reasonable request.

## Declaration of conflicting interests

APyP, QZ and AV report grants from Metabiota Inc, outside the submitted work.

## Notes

**Funding:** This work was supported by the National Institutes of Health R01-AI139761.

### Funding Statement

This work was supported by the National Institutes of Health R01-AI139761.

