## Supplementary File for "Using simulated infectious disease outbreaks to guide the design of individually randomized vaccine trials"

**Figure S1.** Top 100 sites with highest Zika virus transmission probability in the Americas identified by the global epidemic and mobility model (GLEAM) in 2017. Larger circles and darker colors represent greater population size and average site-level incidence of infection, respectively, across all simulated outbreaks in one year.

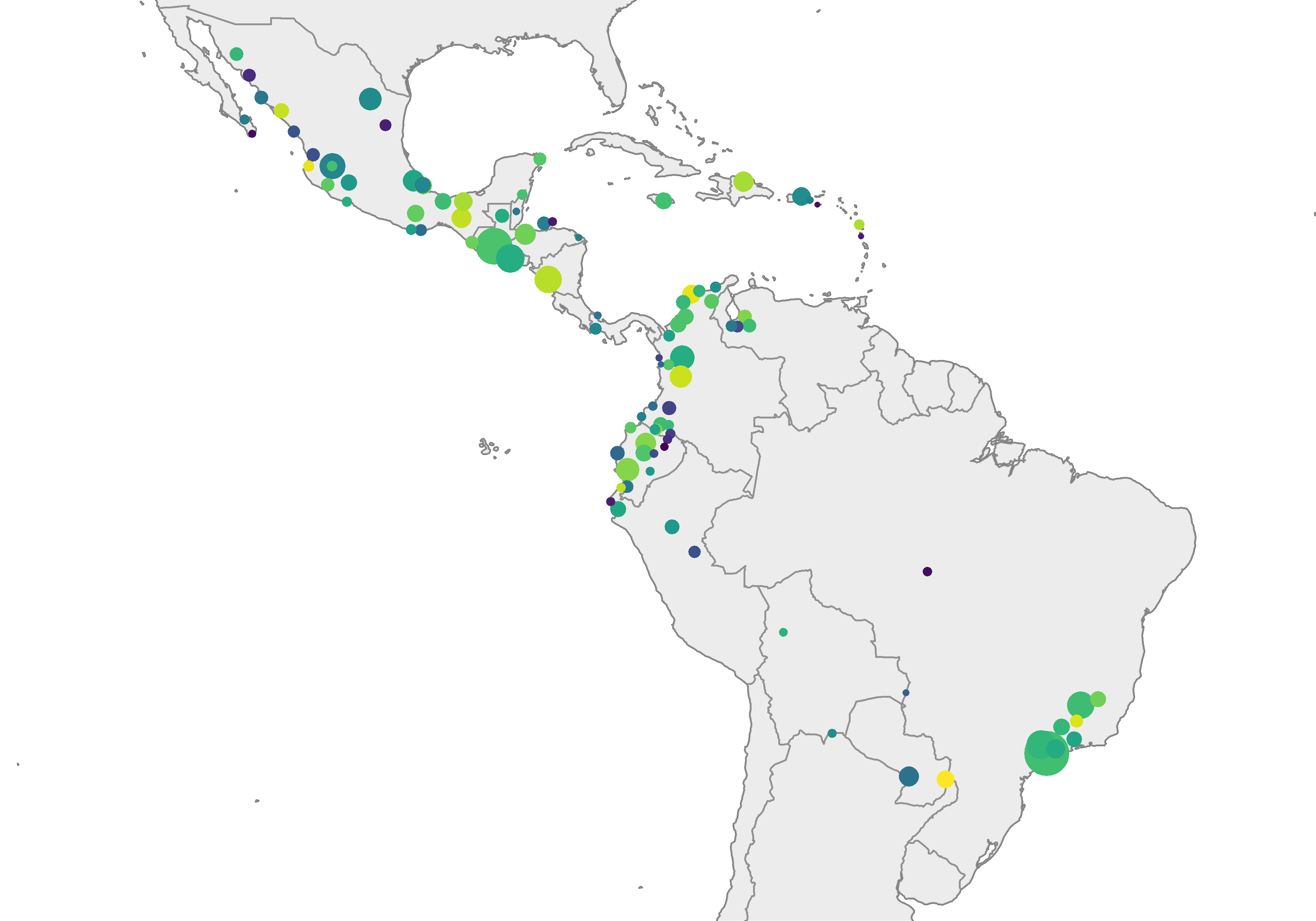

**Figure S2**. Relationships between site prioritization ranking metrics. Each site is plotted, and the color of the dots corresponds to the log site population size. Higher ranks indicate greater 1) median site-level incidence of infection, 2) average site-level incidence, and 3) probability of exceeding 1% site-level incidence across all simulated outbreaks in one year (2017).
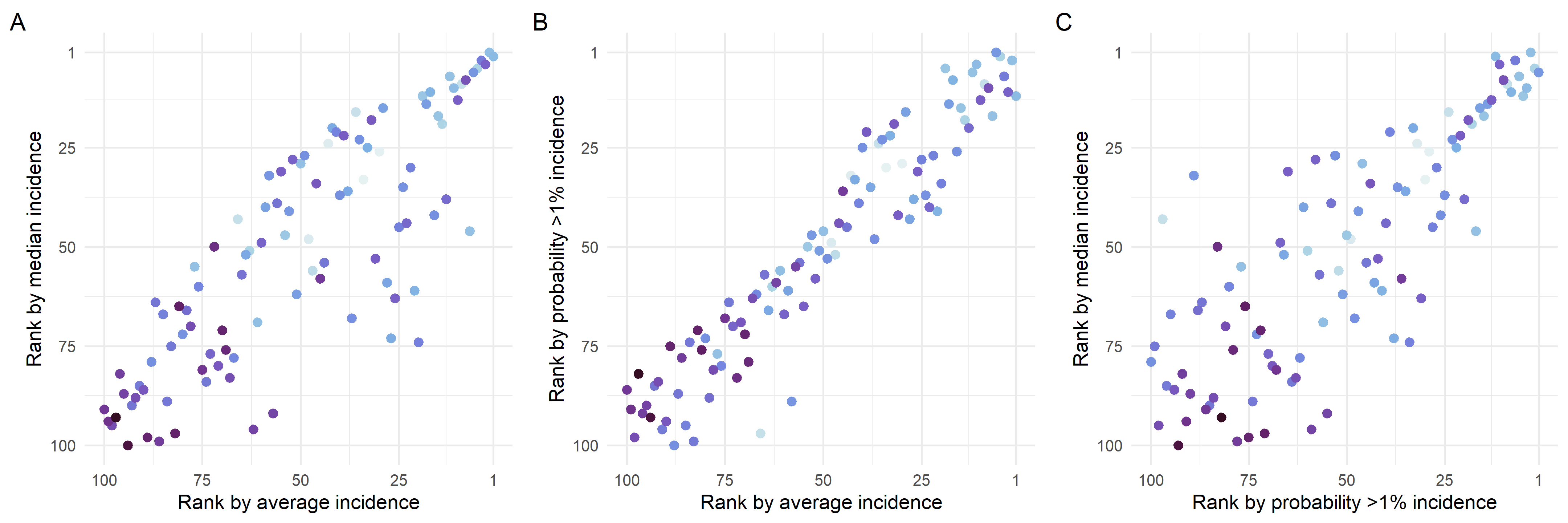

**Figure S3.** Top 100 sites with highest probability of Zika virus transmission in the Americas in 2017 identified by the global and epidemic mobility model (GLEAM) ranked by average site-level incidence of infection across all simulated outbreaks.

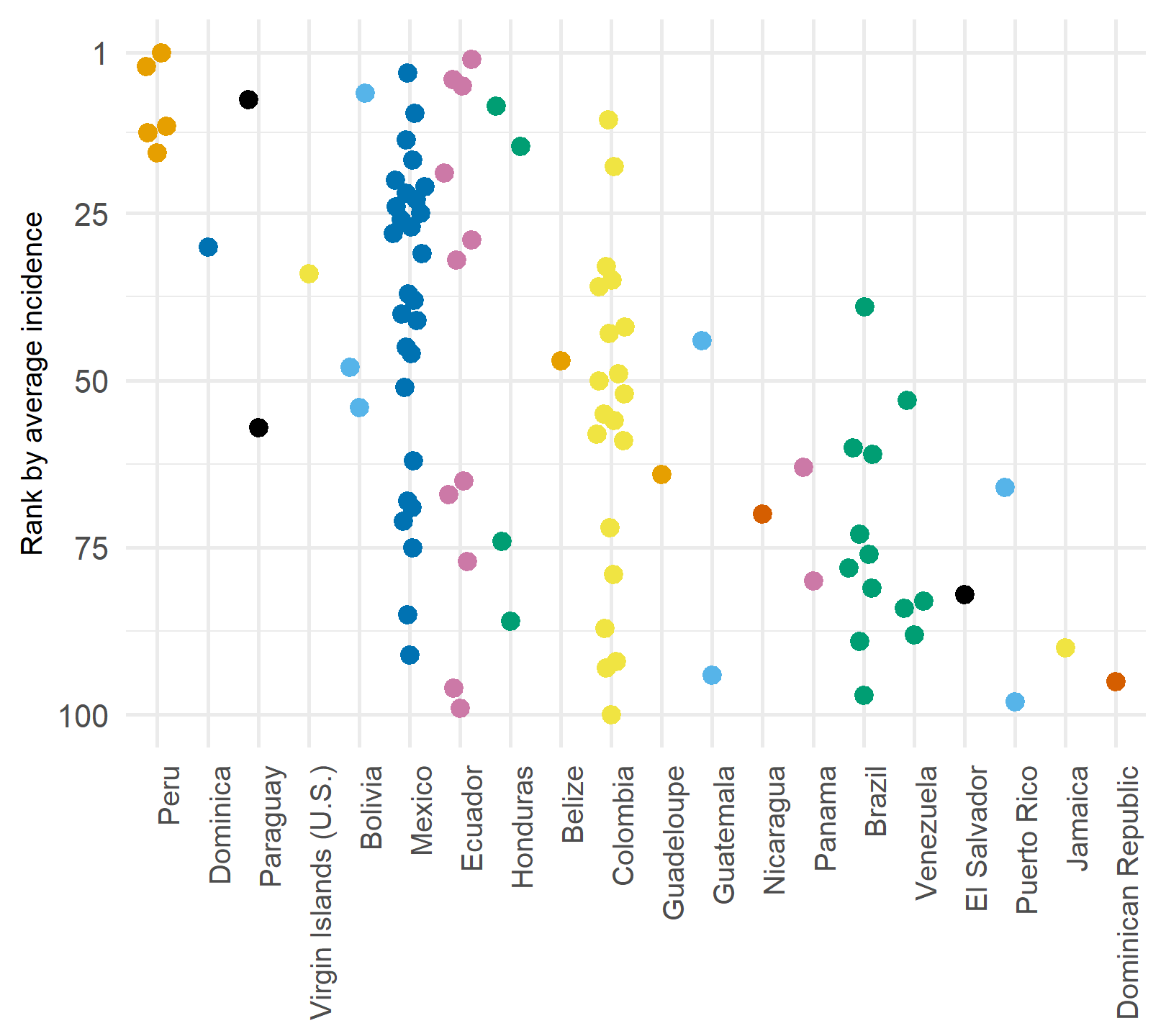

| **Table S1.** Top 100 sites in the Americas with the highest projected Zika virus transmission probability and infection rates in 2017 from the global epidemic and mobility model (GLEAM)^a^ | | |
| --- | --- | --- |
| Site | State | Country |
| Belmopan | Cayo | Belize |
| Rurrenabaque | La Paz | Bolivia |
| Puerto Suarez | Santa Cruz | Bolivia |
| Yacuiba | Tarija | Bolivia |
| Alta Floresta | Mato Grosso | Brazil |
| Belo Horizonte | Minas Gerais | Brazil |
| Ipatinga | Minas Gerais | Brazil |
| Sao Joao del Rei | Minas Gerais | Brazil |
| Varginha | Minas Gerais | Brazil |
| Resende | Rio de Janeiro | Brazil |
| Campinas | Sao Paulo | Brazil |
| Sao Jose Dos Campos | Sao Paulo | Brazil |
| Sao Paulo | Sao Paulo | Brazil |
| Apartado | Antioquia | Colombia |
| Medellin | Antioquia | Colombia |
| Barranquilla | Atlantico | Colombia |
| Cartagena | Bolivar | Colombia |
| Pereira | Caldas | Colombia |
| Popayan | Cauca | Colombia |
| Valledupar | Cesar | Colombia |
| Bahia Solano | Choco | Colombia |
| Nuqui | Choco | Colombia |
| Quibdo | Choco | Colombia |
| Monteria | Cordoba | Colombia |
| Riohacha | La Guajira | Colombia |
| Santa Marta | Magdalena | Colombia |
| Guapi | Nariño | Colombia |
| Ipiales | Nariño | Colombia |
| Pasto | Nariño | Colombia |
| Tumaco | Nariño | Colombia |
| Mocoa | Putumayo | Colombia |
| Puerto Asis | Putumayo | Colombia |
| Corozal (CO) | Sucre | Colombia |
| Dominica | Saint Paul | Dominica |
| Santiago | Santiago | Dominican Republic |
| Macas | Chimborazo | Ecuador |
| Santa Rosa (EC) | El Oro | Ecuador |
| Esmeraldas | Esmeraldas | Ecuador |
| Guayaquil | Guayas | Ecuador |
| Tulcan | Imbabura | Ecuador |
| Manta | Manabi | Ecuador |
| Tena | Napo | Ecuador |
| Coca | Orellana | Ecuador |
| Quito | Pichincha | Ecuador |
| Lago Agrio | Sucumbios | Ecuador |
| Latacunga | Tungurahua | Ecuador |
| San Salvador (SV) | San Salvador | El Salvador |
| Pointe-a-Pitre | Basse-Terre | Guadeloupe |
| Guatemala City | Guatemala | Guatemala |
| Flores | Peten | Guatemala |
| Guanaja | Colon | Honduras |
| San Pedro Sula | Cortes | Honduras |
| Puerto Lempira | Gracias a Dios | Honduras |
| Roatan | Olancho | Honduras |
| Kingston (JM) | Saint Catherine | Jamaica |
| La Paz (MX) | Baja California Sur | Mexico |
| San Jose Cabo | Baja California Sur | Mexico |
| Tapachula | Chiapas | Mexico |
| Tuxtla Gutierrez | Chiapas | Mexico |
| Colima | Colima | Mexico |
| Guadalajara | Jalisco | Mexico |
| Manzanillo (MX) | Jalisco | Mexico |
| Puerto Vallarta | Jalisco | Mexico |
| Lazaro Cardenas | Michoacan | Mexico |
| Uruapan | Michoacan | Mexico |
| Tepic | Nayarit | Mexico |
| Monterrey | Nuevo Leon | Mexico |
| Huatulco | Oaxaca | Mexico |
| Oaxaca | Oaxaca | Mexico |
| Puerto Escondido | Oaxaca | Mexico |
| Cancun | Quintana Roo | Mexico |
| Chetumal | Quintana Roo | Mexico |
| Culiacan | Sinaloa | Mexico |
| Los Mochis | Sinaloa | Mexico |
| Mazatlan | Sinaloa | Mexico |
| Ciudad Obregon | Sonora | Mexico |
| Hermosillo | Sonora | Mexico |
| Villahermosa | Tabasco | Mexico |
| Ciudad Victoria | Tamaulipas | Mexico |
| Jalapa | Veracruz | Mexico |
| Minatitlan | Veracruz | Mexico |
| Tampico | Veracruz | Mexico |
| Veracruz | Veracruz | Mexico |
| Managua | Managua | Nicaragua |
| Bocas del Toro | Bocas del Toro | Panama |
| David | Chiriqui | Panama |
| Ciudad del Este | Alto Parana | Paraguay |
| Asuncion | Central | Paraguay |
| Piura | Piura | Peru |
| Talara | Piura | Peru |
| Tarapoto | San Martin | Peru |
| Tumbes | Tumbes | Peru |
| Pucallpa | Ucayali | Peru |
| Vieques | Fajardo | Puerto Rico |
| San Juan (PR) | Guaynabo | Puerto Rico |
| Barinas | Barinas | Venezuela |
| El Vigia | Merida | Venezuela |
| Merida (VE) | Merida | Venezuela |
| Valera | Trujillo | Venezuela |
| St Croix Island | NA | Virgin Islands (U.S.) |
| ^a^ The site names listed are the largest urban areas associated with the geographic grids used by GLEAM. For example, Belmopan may include areas outside the urban center. | | |

| Table S2. Summary statistics for top ten sites with highest Zika virus transmission probability in 2017 ranked by an equal weight, combined score of the other ranking methods (average site-level incidence of infection, median site-level incidence, probability of exceeding 1% site-level incidence) | | | | | |
| --- | --- | --- | --- | --- | --- |
| Site | State | Country | Average site-level incidence | Median site-level incidence | Probability of >1% of site-level incidence |
| Lago Agrio | Sucumbios | Ecuador | 0.094 | 0.097 | 0.903 |
| Coca | Orellana | Ecuador | 0.063 | 0.060 | 0.905 |
| Esmeraldas | Esmeraldas | Ecuador | 0.060 | 0.059 | 0.943 |
| Los Mochis | Sinaloa | Mexico | 0.080 | 0.071 | 0.741 |
| Tumbes | Tumbes | Peru | 0.130 | 0.073 | 0.613 |
| Piura | Piura | Peru | 0.093 | 0.070 | 0.623 |
| Talara | Piura | Peru | 0.038 | 0.040 | 0.754 |
| Tumaco | Narino | Colombia | 0.039 | 0.026 | 0.780 |
| Ciudad del Este | Alto Parana | Paraguay | 0.053 | 0.030 | 0.667 |
| Puerto Lempira | Gracias a Dios | Honduras | 0.048 | 0.029 | 0.701 |

| **Table S3.** Example of the number of participants enrolled from each of five sites for different enrollment strategies (n = 15,000) | | | |
| --- | --- | --- | --- |
| Site Rank^a^ | Equal across sites | Proportional to average incidence | (Equal + average incidence)/2 |
| 1 | 3000 | 4242 | 3621 |
| 2 | 3000 | 3059 | 3030 |
| 3 | 3000 | 3032 | 3016 |
| 4 | 3000 | 2606 | 2803 |
| 5 | 3000 | 2061 | 2530 |
| ^a^ Ranked by average site-level incidence of infection across simulated outbreaks. | | | |
